# Genetic association of T1D stratified by HLA DR3 and DR4 status reveals heterogeneity in pathways of progression to T1D

**DOI:** 10.1101/2025.08.13.25333386

**Authors:** Amber M Luckett, Carolyn McGrail, KA Murrall, Emily N Griffin, Robin N Beaumont, Gareth Hawkes, William A Hagopian, Stephen S Rich, Michael N Weedon, Sarah Richardson, Richard A Oram, Kyle J Gaulton

## Abstract

**Background:** There are substantial differences in the clinical presentation of type 1 diabetes (T1D) depending on the first-developing autoantibody, although the underlying mechanisms are poorly understood. The *DR3-DQ2* (DR3) and *DR4-DQ8* (DR4) haplotypes at the MHC locus closely associate with GAD and IAA as the first-developing autoantibody, respectively, and can therefore be used as proxies for first autoantibody development in large cohort studies of T1D cases and controls.

**Methods:** We performed the first genome-wide association study of T1D stratified by DR3 and DR4 status using 9,091 T1D cases and 14,157 controls from multiple cohorts. We estimated heritability and genetic correlation between DR3-T1D and DR4-T1D, and with other immune and glycaemic phenotypes. We assessed heterogeneity in effects on T1D between DR3 and DR4 individuals at known T1D loci. We determined enrichment of T1D heritability in DR3 and DR4 among variants in cell type-specific *cis*-regulatory elements (cREs) and biological pathways, and annotated risk variants in cREs.

**Results:** We observed only moderate genetic correlation between DR4- and DR3-T1D (rg=0.6), which was lower compared to stratifications based on age of onset and sex, and distinct patterns of genetic correlations with other autoimmune diseases. Among T1D-associated loci, the *IL2* locus had significantly larger effect on T1D in DR4 while several other loci (*TAGAP, KLRG1*) had more nominal heterogeneity. There was stronger enrichment of DR4-T1D associated variants in T cell cREs and T cell-related pathways, while DR3-T1D associated variants were specifically enriched in mast cell cREs. We finally prioritized specific loci annotated in mast cells with stronger effects on T1D in DR3 individuals.

**Conclusion:** We performed the first GWAS of T1D stratified by DR3 and DR4 status, which revealed heterogeneity in genetic risk and biological mechanisms dependent on high-risk HLA background.

## INTRODUCTION

Type 1 diabetes (T1D) is a complex disease characterised by autoimmune destruction of beta cells, but the underlying aetiology is not well understood [1]. The timing of progression to clinical diabetes varies widely across individuals, ranging from very early in childhood to adult onset [2, 3]. The development of autoantibodies (AAB) against islet proteins such as GADA (glutamic acid decarboxylase) and IAA (insulin) is a hallmark of disease progression in many T1D cases. Recent studies have argued there are distinct endotypes of T1D marked by the first-appearing AAB, where GADA-first T1D typically has later onset compared to IAA-first T1D [2, 3]. Insufficient data from cohort studies and the transient nature of AAB, however, have prohibited a deeper understanding of potential heterogeneity in disease processes leading to the development of GADA-first and IAA-first T1D [2, 4].

The major histocompatibility complex (MHC) encodes human leukocyte antigen (HLA) genes vital in adaptive and innate immune responses [5], and the MHC locus confers substantial T1D risk in European ancestry individuals. The largest T1D risk factors at the MHC locus include the HLA haplotypes *DRB1*^*\**^*03:01–DQA1*^*\**^*05:01–DQB1*^*\**^*02:01* (*DR3-DQ2*, denoted DR3) and *DRB1*^*\**^*04:XX– DQA1*^*\**^*03:01–DQB1*^*\**^*03:02* (*DR4-DQ8*, denoted DR4) [5–8]. There is evidence to suggest a relationship between the presence of these HLA haplotypes and the first-developed AAB in individuals who develop T1D, where DR3 is associated with GAD-first AAB and later disease onset and DR4 is associated with IAA-first AAB and earlier disease onset [2]. In addition, individuals with DR4 are more responsive to preventative T1D treatments targeting T cell responses such as methyldopa and teplizumab, which supports that distinct processes impact T1D in different HLA backgrounds [4, 9, 10].

We hypothesized that identifying differences in genome-wide genetic risk of T1D in individuals with DR3 or DR4 would help reveal novel insight into differing processes involved in T1D progression related to first-developing AAB. Large T1D case and control cohorts with genetic data enable performing genetic association studies of T1D stratified by DR3 or DR4 as a proxy for first-developed autoantibody where cohort sizes are much smaller. We therefore performed a genome-wide association study of T1D in individuals stratified by DR3 and DR4 and characterized heterogeneity in T1D genetic risk and genomic mechanisms.

## METHODS

### Study cohorts and genotype imputation

We compiled genotype data from 17,413 T1D and 37,570 control individuals of European ancestry from publicly available cohorts (dbGaP) and from Exeter, United Kingdom **(Supplementary Table 1)**. European ancestry individuals were used in this analysis due to the higher frequency of DR3 and DR4. Individuals with first four PCs >3 interquartile ranges from the 25th and 75th percentiles of PCs from European 1000 Genomes Project [11] were considered non-European and excluded. T1D case cohorts were matched to control cohorts consistent with country of origin and genotype array, as previously described [12, 13]. Prior to imputation, we applied the HRC imputation preparation program (v4.2.9, https://www.well.ox.ac.uk/~wrayner/tools/) and used PLINK (v1.90) to perform quality control to remove variants with a difference in allele frequency >0.2 compared to the HRC r1.1 reference panel. We then removed variants with MAF <1%, missing genotypes >5%, in violation of Hardy-Weinberg equilibrium (HWE *P*<1×10-5 in control cohorts and HWE *P*<1×10-10 in case cohorts) and variants with allele ambiguity. We removed samples with missing genotypes >5%, sex mismatch based on reported phenotypes, non-European ancestry, and cryptic relatedness through identity-by-descent (IBD>0.2).

We used the TOPMed Imputation Server r2 panel and Michigan imputation Server multi-ethnic HLA reference panel for all samples **(Figure 1A)** [14, 15]. For genome-wide imputation, we removed variants with an imputation accuracy score of r^2^<0.3. For HLA variant imputation, we used a threshold of r^2^<0.5 to remove variants. Variants which had a standard deviation in control allele frequency >0.055 across cohorts were also removed. Variants that passed QC filters in all cohorts were tested for association.

**Figure 1.**
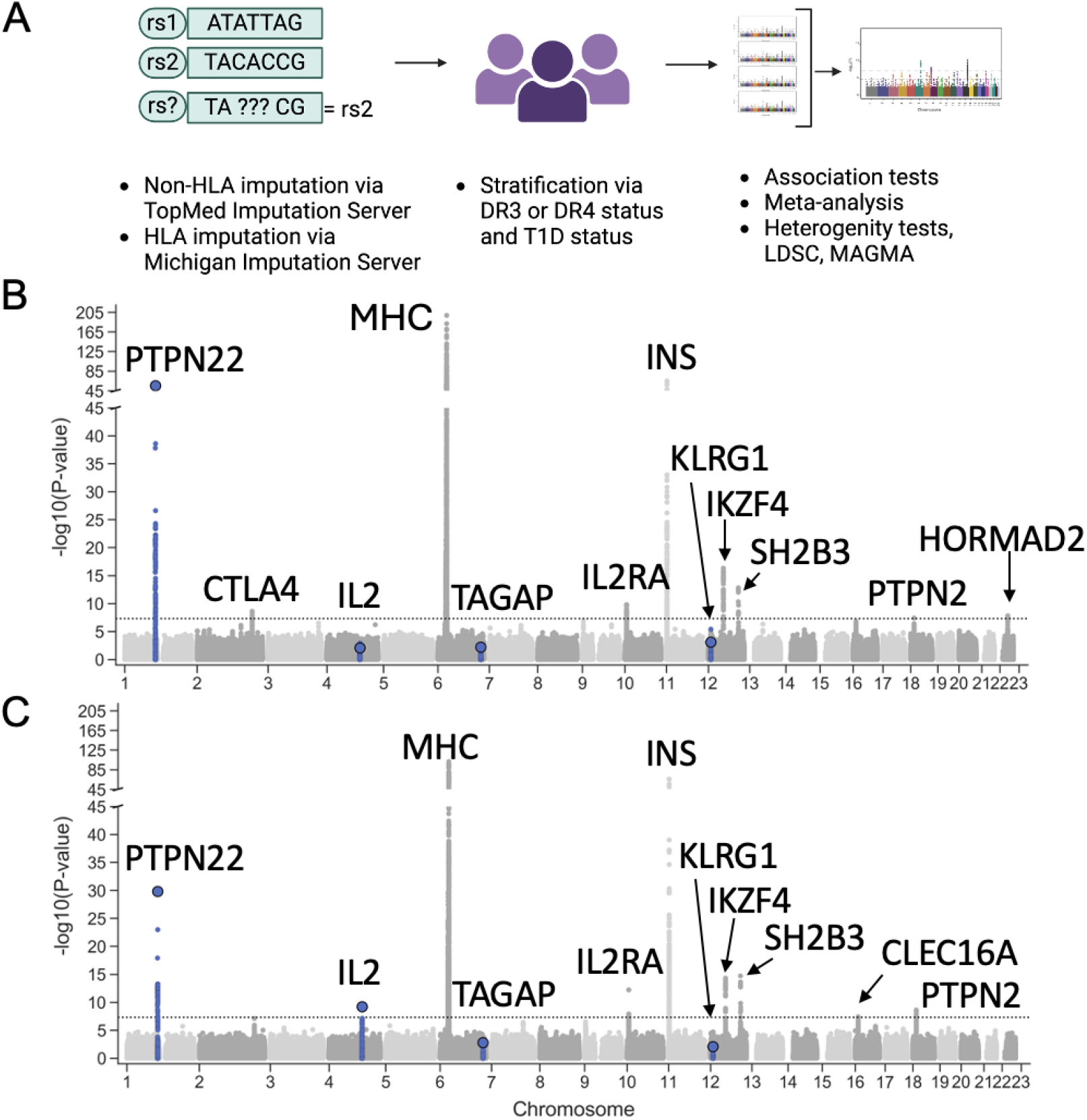
Genetic discovery for T1D stratified by HLA-DR3 and DR4. (A) Overview of genetic asociation and heterogeneity tests for T1D in DR3 and DR4 individuals. (B) Genome-wide T1D association in individuals with a *DR3-DQ2* haplotype. log_10_ *P* values from meta-analysis of 4057 T1D cases and 8043 non-T1D controls. (C) Genome-wide T1D association in individuals with a *DR4-DQ8* haplotype. (log_10_ *P* values from meta-analysis of 5034 T1D cases and 6114 non-T1D controls. For both plots, significant loci are labelled with the nearest gene and loci with heterogeneous effects are colored blue. Dotted line indicates genome-wide significance.

We defined distinct DR3 and DR4 groups based on *DR* and *DQ* haplotype status using four-digit HLA alleles imputed from the Michigan Imputation Server. DR3 was identified by the presence of HLA *DRB1*^*\**^*03:01-DQB1*^*\**^*02:01*, while DR4 was identified by the presence of HLA *DRB1*^*\**^*04:01/02/04/05/08-DQB1*^*\**^*03:02/04*. We partitioned individuals by HLA *DR* and *DQ* haplotype resulting in 4057 DR3-T1D cases and 8043 DR3 controls and 5034 DR4-T1D cases and 6114 DR4 controls **(Supplementary Table 1 and Figure 1A)**.

### Association testing and meta-analysis

For dbGaP cohorts, we tested TOPMed imputed variants for T1D case and control association using Firth bias corrected logistic regression in EPACTS in both DR3 and DR4 groups. We tested variants with MAF>1% for association using sex and the first 4 genotype PCs as covariates. For Exeter cohorts which have smaller sample sizes, we used REGENIE (https://rgcgithub.github.io/regenie/), which is suited to dealing with smaller cohorts by fitting a regression model to a subset of genetic markers prior to trait association testing using Firth bias corrected logistic regression [16]. We used variants with MAF>1% to generate LOCO (leave one chromosome out) predictions and used sex and the first 4 genotype PCs as covariates. We then performed association analysis in each cohort.

Summary statistics were combined across cohorts in a fixed effects inverse variance weighted meta-analysis. We removed HLA variants and calculated genomic inflation (λgc) for the DR3-T1D GWAS (λgc= 1.02) and DR4-T1D GWAS (λgc = 1.0).

### Heritability and genetic correlation estimates

Using LD Score Regression (LDSC) we estimated the heritability of DR3-T1D and DR4-T1D, including and excluding the HLA region, using the liability scale [17]. We used a population prevalence of 0.0012 and a sample prevalence of 0.34 for DR3-T1D and 0.45 for DR4-T1D. We then performed genetic correlation estimation between DR3-T1D and DR4-T1D, and within females with T1D vs males with T1D, and younger vs older onset T1D (the lower median vs higher median T1D age of onset from T1DGC, where the median was 8 years), as a positive control comparison due to high genetic correlation between these stratifications.

To determine genetic similarity between DR3-T1D, DR4-T1D and other traits, we used published GWAS summary statistics to estimate heritability, and genetic correlation estimates for 10 autoimmune traits (autoimmune thyroid disease, coeliac, Crohn’s, multiple sclerosis, primary biliary cirrhosis, primary sclerosing cholangitis, rheumatoid arthritis, systemic lupus erythematosus, T1D, ulcerative colitis, and vitiligo) and 3 non-autoimmune traits (type 2 diabetes, BMI, and pancreas volume) [12, 18–28]. Sample and population prevalences were calculated from each GWAS and from current literature, respectively. We used Benjamini-Hochberg correction to determine whether correlations were significant (FDR<0.10).

### T1D variant heterogeneity testing between HLA stratified groups

We tested heterogeneity in lead variants with MAF >1% from previously described T1D fine-mapped signals [12] using a merged matrix of genotypes from dbGaP cohorts. We used PLINK to perform a Firth regression heterogeneity test (excluding MHC locus) to examine heterogeneity between DR3-T1D and DR4-T1D within these established T1D signals. For this interaction test, we included the covariates from logistic regression (PCs 1-4, and sex). We then performed a sign test to determine whether there were more extreme differences in effect in either DR3-T1D or DR4-T1D from all 87 T1D variants.

### GWAS enrichment analysis

In the dbGaP cohorts, we used LDSC to perform a partitioned heritability analysis to estimate enrichment of T1D heritability in cell type-specific *cis*-regulatory elements (cREs) defined by CATlas [29]. We used the summary statistics for each group excluding the MHC locus and formatted it for input to LD score regression. We computed cell specific LD enrichment scores for DR3- and DR4-T1D using the version 2.2 1000 Genomes baseline model and corrected for multiple tests (Benjamini Hochberg correction) across all cell types using false discovery rate (FDR). FDR<0.10 was considered significant. We used MAGMA (with default parameters) to perform gene set enrichment analysis in GO, KEGG and REACTOME pathways with DR3- and DR4-T1D summary statistics.

### Mast Cell Specific Gene Heterogeneity

From previously determined 99% credible sets for 136 T1D signals, we identified known T1D signals and determined overlaps within mast cell cREs defined by CATlas [29]. We then determined which variants were present in the meta-analyses for both DR3- and DR4-T1D. We removed variants with a PIP<0.1 as these are less likely to be causal within the credible set. We then examined differences in strength of association to T1D between the stratified HLA groups in credible sets that fell within accessible chromatin of mast cells.

### Ethics

Ethics for the DARE study was granted by the Devon & Torbay Research Ethics Committee, ref: 2002/7/118. The StartRight study was approved by the Southwest–Cornwall and Plymouth NHS Research Ethics Committee (reference: 16/SW/0130). The Extremely Early-Onset Type 1 Diabetes (EXE-T1D) study has ethical approval from Derby Research Ethics Committee, Derby, U.K. (IRAS project ID 228082). The EXTEND/PRB study has ethical approval from Southwest-Cornwall & Plymouth NHS Research Ethics Committee, Bristol, U.K. (reference 14/SW/1089; 5-year extension following initial approval: reference 09/H0106/75). The TIGI study was approved by the Southwest-Central Bristol Research Ethics Committee (reference 13/SW/0312). The Institutional Review Board of the University of California San Diego gave ethical approval to analyse de-identified cohort data obtained from public sources such as dbGAP and EGA.

## RESULTS

### Genome-wide association study of T1D stratified by DR3/DR4 status

We obtained microarray genotype data of 17,413 T1D case and 37,570 control European ancestry samples from 11 cohorts, and imputed genotypes into 308 million and 56,310 variants in the TOPMed v3 and Michigan HLA reference panels, respectively. We then stratified samples based on the presence of DR3 and DR4 haplotypes (**see Methods**), resulting in 12,100 DR3-T1D and control samples and 11,148 DR4-T1D and control samples. For these analyses we excluded individuals who were heterozygous DR3/DR4, as they could not be uniquely assigned to one group. We then tested variants genome-wide for T1D case and control association separately within the DR3 and DR4 groups (**Figure 1A**).

In total, variants at 11 loci reached genome-wide significance (*P*<5×10^-8^) in either the DR3- or DR4-T1D association analysis, all of which were loci previously reported in T1D risk (**Figure 1B-C**). Of these 11 loci, seven (*PTPN22, IL2RA, INS, IKZF4, SH2B3, PTPN2*, MHC) reached genome-wide significance in both analyses (**Figure 1B-C**). Four additional loci reached genome wide significance for T1D in only one analysis, including *CTLA4* and *HORMAD2* in DR3 individuals and *CLEC16A* and *IL2* in DR4 individuals (**Figure 1B-C**).

### Heritability and genetic correlation estimates

We next calculated heritability estimates for DR3-T1D and DR4-T1D including and excluding the HLA region. The overall heritability for DR3-T1D (h^2^= 0.45, SE= 0.06) was higher but not statistically different (p=0.59) to the DR4-T1D heritability (h^2^= 0.40, SE= 0.07) **(Figure 2A)**. After excluding the HLA region, both heritability estimates were as expected significantly reduced *(P*<0.05) with DR3-T1D having higher heritability (h^2^=0.17, SE=0.03) than DR4-T1D (h^2^=0.14, SE=0.03), though again not statistically significant (p=0.39) **(Figure 2A)**.

**Figure 2.**
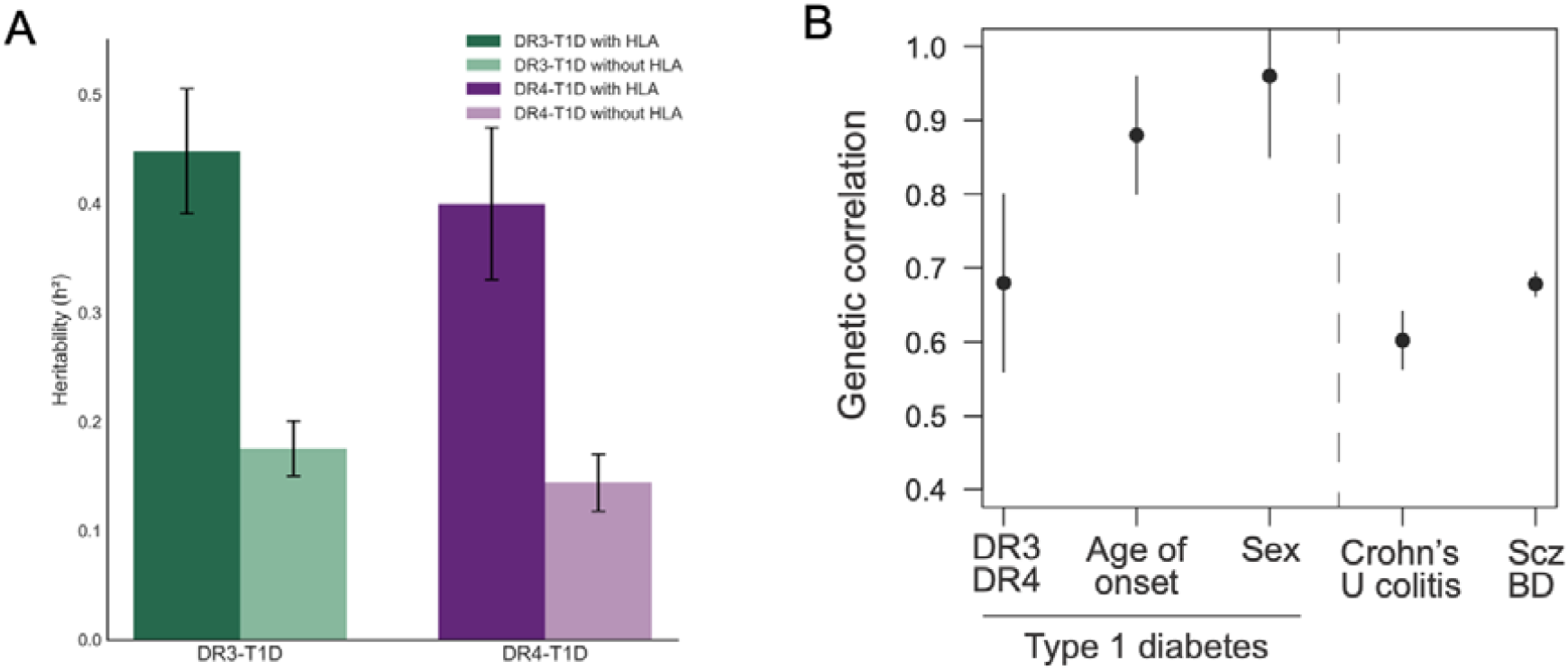
Heritability and Genetic Correlations in DR3 and DR4 T1D. (A) Heritability estimates for DR3-T1D, including HLA variants (dark green) and excluding HLA variants (light green), and DR4-T1D including HLA variants (dark purple) and excluding HLA variants (light purple) with 95% confidence intervals as error bars. (B) Genetic correlation in T1D association stratified by DR3 and DR4, median age of onset (diagnosis), and biological sex with 95% confidence intervals as error bars. For comparison the genetic correlation between Schizophrenia and Bipolar disorder, and Crohn’s disease and Ulcerative colitis are also shown.

We next determined the correlation in genome-wide genetic effects on T1D in DR3 and DR4 individuals. After excluding the HLA region, we found that DR3-T1D and DR4-T1D was moderately correlated (rg= 0.68, SE=0.12) in genome-wide genetic effects. By comparison, we observed higher correlation among T1D association between females and males (rg=0.88, SE=0.08), as well as in younger age of onset (<8 years based on T1DGC median) vs older age of onset T1D (≥8 years based on T1DGC median) (rg= 0.96, SE=0.11) **(Figure 2B)**. The correlation between DR3- and DR4-T1D was significantly lower than the correlation between sexes and age of onset (P<.05), and similar to the correlation between Crohn’s disease and Ulcerative colitis and Schizophrenia and Bipolar disorder. These results demonstrate that T1D in DR3 and DR4 individuals shows less genetic similarity compared to previously stratifications of T1D individuals.

When assessing genetic correlation with other autoimmune and endocrine traits (excluding the HLA region), we found both shared and specific correlations between DR3-T1D or DR4-T1D and other traits **(Supplementary Figure 1A-B)**. After multiple test correction, both DR3-T1D and DR4-T1D were significantly (FDR<0.10) positively genetically correlated with rheumatoid arthritis (DR3-T1D rg=0.40, DR4-T1D rg=0.38), autoimmune thyroid disease (DR3-T1D rg=0.37, DR4-T1D rg=0.37), and systemic lupus erythematosus although the correlation was stronger in DR4-T1D (DR3-T1D rg=0.19, DR4-T1D rg=0.41) **(Supplementary Figure 1B)**. Among traits with more specific enrichments, primary biliary cirrhosis was significantly (FDR<0.10) positively correlated with DR3-T1D only (DR3-T1D rg=0.22, DR4-T1D rg=0.12) and coeliac disease was significantly positively correlated with DR4-T1D only (DR4-T1D rg=0.3; DR3-T1D rg=0.05) as was vitiligo (DR4-T1D rg=0.31, DR3-T1D rg=0.16) **(Supplementary Figure 1B)**.

### Heterogeneity in variant effects on T1D in DR3 and DR4 individuals

We next examined heterogeneity in the effects of variants at specific risk loci on T1D risk between DR3- and DR4-T1D. First, we determined whether the lead variants at the 10 non-MHC loci with genome-wide significant association had heterogeneous effects on T1D using an interaction test (**see Methods**). At nominal significance (P<0.05), the *PTPN22* locus had a stronger effect in DR3-T1D (DR3-T1D beta= 0.738, DR4-T1D beta=0.604, *P*=0.03) whilst the *IL2* locus had a stronger effect in DR4-T1D (DR3-T1D beta=0.0878, DR4-T1D beta=0.230, *P*=0.006). After multiple test correction (FDR<0.10), only *IL2* had evidence for significant heterogeneity in effects between DR3- and DR4-T1D (**Figure 3A-B**).

**Fig 3.**
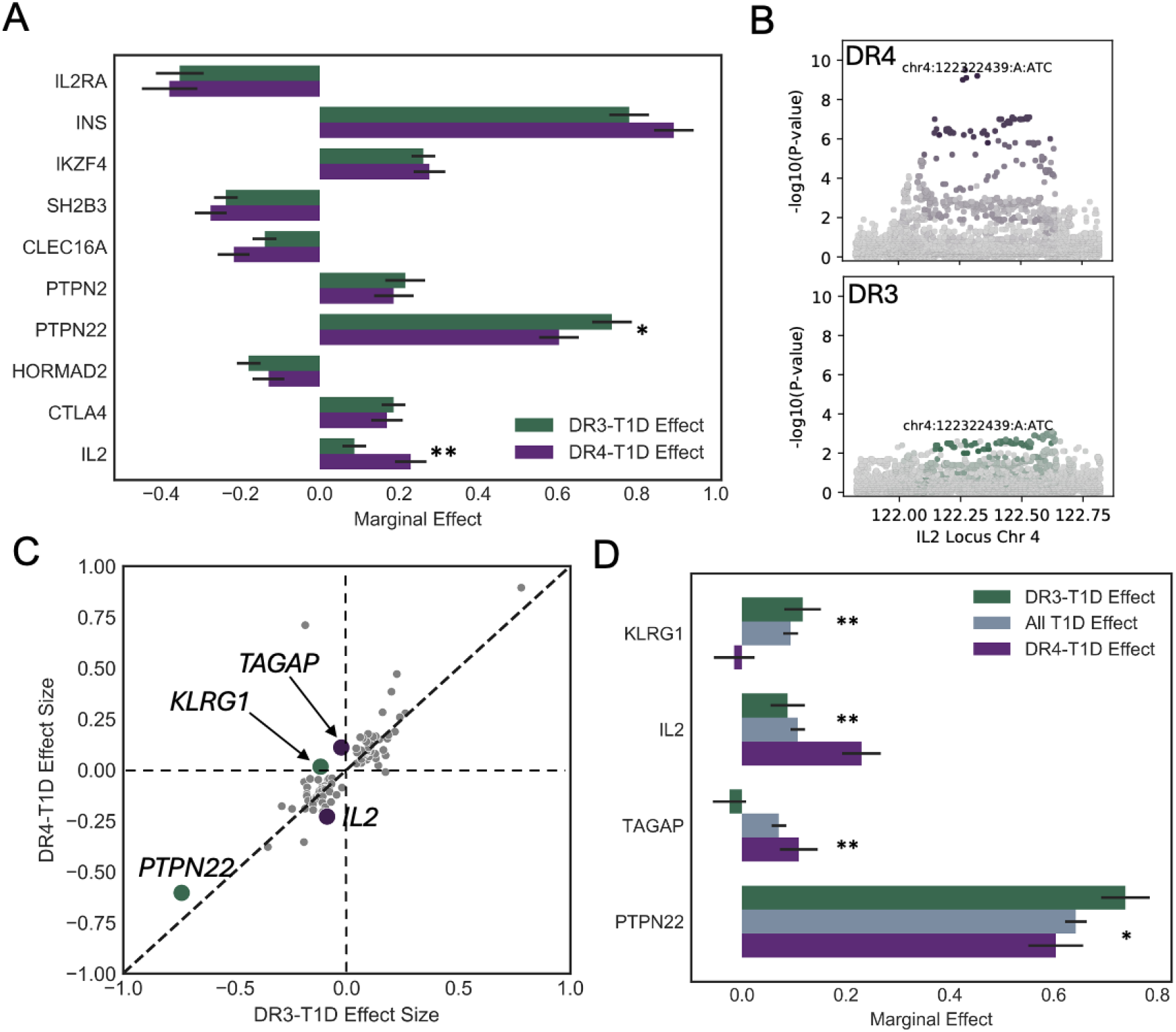
Genetic Heterogeneity between T1D in DR3 and DR4. (A) Plot of effect sizes of genome-wide significant loci in DR3-T1D (green) and DR4-T1D (purple). (B) T1D association at the *IL2* locus in DR4 (top) and DR3 (bottom) individuals. (C) Genetic effects on T1D in DR3 and DR4 at 87 known T1D loci. Larger points indicate signals with evidence for heterogeneity (*P*<0.05). Purple points indicate stronger effect size in DR4-T1D. Green points indicate stronger effect size in DR3-T1D. D. Plot of effect sizes for heterogenous signals genome-wide between DR3-T1D (green) and DR4-T1D (purple) with overall T1D effect sizes as a comparison. *P*<0.05=^*^, *P*<0.01=^**^. All error bars are standard errors.

Next, we more broadly determined whether lead variants across known T1D risk loci outside of these 10 significant loci had significant heterogeneity between DR3-T1D and DR4-T1D. We tested the heterogeneity of an additional 77 T1D signals and found no additional loci with significant heterogeneity after FDR correction, although we found an additional two loci with nominal evidence (uncorrected *P*<0.05) (**Figure 3C-E)**. The *TAGAP* locus had a larger effect in DR4-T1D (DR3-T1D beta=-0.024, DR4-T1D beta=0.11, *P*=0.0077) and the *KLRG1* locus had a stronger effect in DR3-T1D (DR3-T1D beta=0.11, DR4-T1D beta=-0.015, *P*=0.0050). This heterogeneity is also reflected in stratified association results at these loci **(Supplementary Figure 2)**.

Finally, we determined whether there was evidence for pronounced effect of variants on T1D risk broadly between DR3-T1D and DR4-T1D individuals. A sign test between the 87 T1D risk loci lead variant effects from DR3-T1D and DR4-T1D yielded no significant difference between groups (p=0.67).

### Cell type-specific enrichment for T1D association in DR3 and DR4

Given evidence for heterogeneity in genetic effects on T1D between DR3-T1D and DR4-T1D individuals, we next sought to understand the biological mechanisms underlying these differences.

We first performed enrichment analyses of DR3-T1D and DR4-T1D association for *cis*-regulatory elements (cREs) active in immune cell populations using LD score regression and identified immune cell types with significant enrichment (FDR<0.10) in each group (**Supplementary Table 2)**. In both the DR3-T1D and DR4-T1D group, we observed significant enrichment (FDR<0.10) of T1D associated variants in CD4+ T-cell and CD8+ T-cell cREs with DR4-T1D exhibiting stronger enrichment in each of these cell types (**Figure 4A**). By comparison, the DR3-T1D group had a stronger enrichment of T1D associated variants in foetal thymocyte, foetal cytotoxic T-cell and foetal CD4+ T-cell cREs (**Figure 4A**). Interestingly, we also observed highly specific enrichment of T1D associated variants in mast cell cREs for DR3-T1D not observed in the DR4-T1D population.

**Fig 4.**
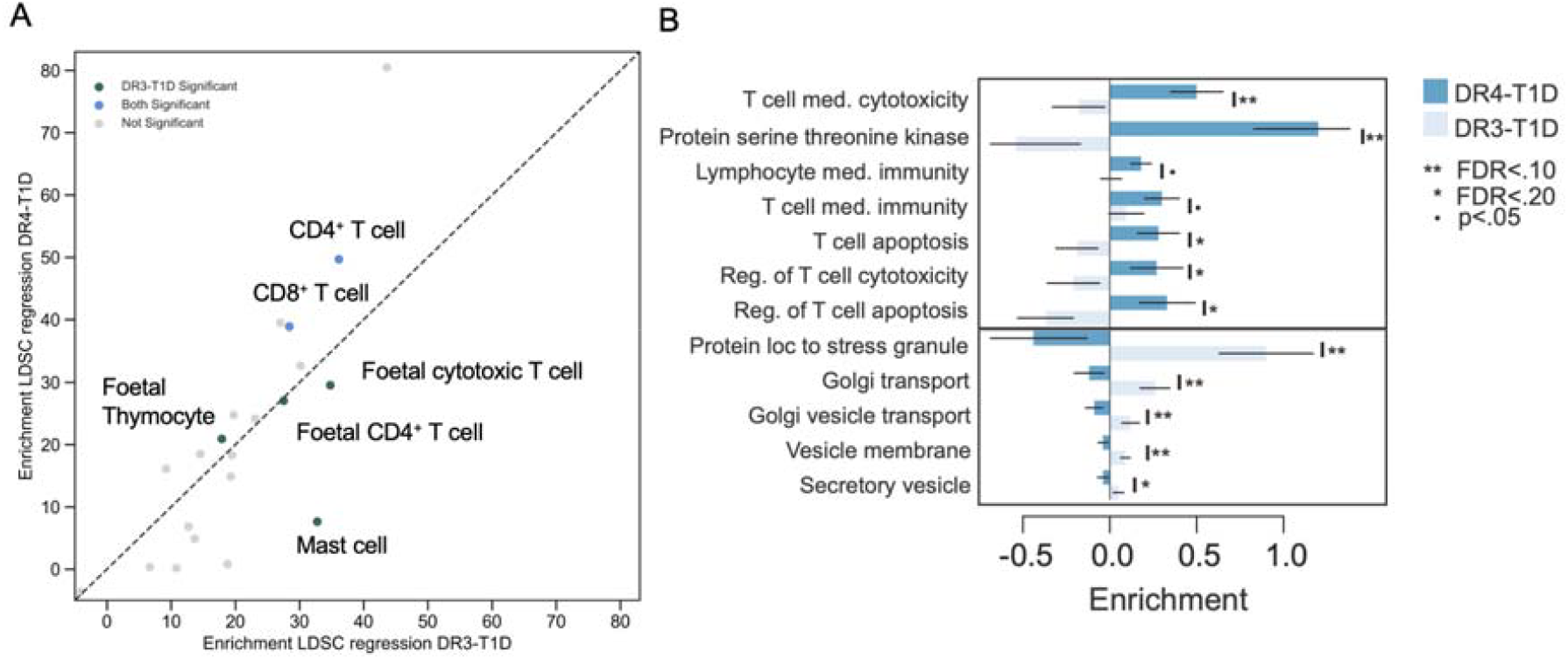
Functional annotations enriched in T1D association in DR3 and DR4. (A) Enrichment scores of T1D-associated variants in DR3 and DR4 for stimulated and unstimulated immune cell accessible chromatin sites using LD score regression. Blue points are significant at FDR<0.10 in both DR3-T1D and DR4-T1D while green points are FDR significant in only DR3-T1D, and grey points are not significant. (B) Enrichment of biological pathways for T1D association in DR3 and DR4 using MAGMA. Blue bars are enrichment scores in DR4-T1D xand light blue are enrichment scores in DR3-T1D. ^**^FDR<.10, ^*^FDR<.20, •p<.05

We next identified biological pathways enriched for DR3- and DR4-T1D associated variants using MAGMA. Among pathways enriched in either DR3- or DR4-T1D, we further identified pathways with heterogeneity (FDR<.20) in enrichment estimates between the two groups (**Figure 4B, Supplementary Table 3**). Pathways with more pronounced enrichment in DR4-T1D included multiple related to T-cell activity, signalling and apoptosis and immune responses. By comparison, pathways with stronger enrichment (FDR<.20) in DR3-T1D were highly distinct and included those related to secretory vesicles, stress granules, and several other processes. This reveals differences in biological pathways enriched for T1D association in DR3 and DR4 individuals.

T1D associated variants showed highly specific enrichment in mast cell cREs in DR3-T1D individuals and not DR4-T1D, suggesting mechanistic differences in cell types involved in disease progression, which we investigated further. We first identified T1D associated loci with candidate causal variants overlapping mast cell cREs. In total, seven T1D loci contained at least one candidate variant (PIP>0.1) in a mast cell cRE, five of which had stronger effects in DR3-T1D compared to DR4-T1D (**Supplementary Table 4**). Among these were variants at the *FLI1* locus, where the *FLI1* gene has been implicated in histamine production in mast cells [30]. Next, we identified variants with sub-genome-wide significant T1D association in mast cell cREs. Among loci at more nominal thresholds were several which had heterogeneous effects on T1D between DR3- and DR4-T1D and had no evidence for T1D association in DR4. For example, variants at the 10q24 locus showed evidence for T1D association in DR3 (beta=-0.13, p=5.7×10^-5^) but not in DR4 (beta=0.001, p=0.97) (**Supplementary Figure 3**). These variants mapped in the *SUFU* gene, which has been implicated in regulating several key transcription factors in mast cells [31].

## DISCUSSION

Genetic association of T1D in individuals stratified by HLA DR3 and DR4 haplotypes revealed evidence for heterogeneity in disease risk dependent on high-risk HLA background. The genetic correlation between DR3- and DR4-T1D was significantly lower relative to other stratifications of T1D such as sex or age of onset. Furthermore, the correlation between DR3- and DR4-T1D was comparable to correlations between different forms of other complex diseases such as psychiatric disease (schizophrenia and bipolar disorder) [32] and inflammatory bowel disease (Crohn’s disease and Ulcerative colitis) [33]. In addition to broad heterogeneity in T1D risk, we identified specific loci with heterogeneous effects between DR3 and DR4 mostly prominently *IL2* which had more pronounced effects in DR4. Together this reveals that T1D in a DR3- and DR4 background may represent heterogeneous sub-forms of T1D at the genetic level, which is line with the observed clinical differences in T1D presentation dependent on first-appearing autoantibody.

We also identified distinct enrichments among T1D-associated variants in DR3- and DR4-T1D for regulatory elements in immune cell types, highlighting potential differences in underlying disease mechanisms. Mast cells were specifically enriched for T1D associated variants in DR3-T1D, which has not been described previously in T1D GWAS possibly due to a lack of DR4-T1D enrichment masking the effect. Mast cell activity may exacerbate T1D progression in response to environmental exposures in DR3-T1D [34]. Altered mast cell activity in T1D may thus have more temporal or spatial components, in addition to possible differences in gene expression and not overall numbers [35, 36]. Our findings also provide further evidence of greater T-cell response and enrichment in DR4-T1D. We also observed heterogeneity with increased effect in DR4-T1D at *IL2* which plays key roles in T-cell activation and differentiation [37, 38]. The more pronounced role of T cells in DR4-T1D compared to DR3-T1D may guide personalised medicine using methyldopa, which binds to HLA-*DQ8* to reduce T-cell responses [10], and teplizumab, which binds to autoreactive T cells resulting in DR4 carriers responding better in absence of DR3 [9].

We utilized DR3 and DR4 status as a proxy for first-appearing antibody, although there isn’t a perfect correlation between first-appearing GAD and IAA and HLA DR3 and DR4 alleles (2). Future T1D GWAS of individuals with first-appearing AAB directly will be needed to further establish the differences in T1D risk driven by AAB status. A current limitation, however, is the small available sample sizes of individuals where both first-developing AAB and genotyping data is available. In addition, there may be differences between the heterozygous combination of DR3 and DR4 compared to homozygous or single carriers of each haplotype. Heterozygous DR3/DR4 conveys the highest T1D risk in European ancestry [7], however the association of heterozygous DR3/DR4 with first-developing AAB is unclear and the high T1D risk of this haplotype combination means the control group sample size is limited [2]. In addition, the transient nature of T1D AAB detection means prospective cohort studies like TEDDY [39] and TrialNET [40], which monitor T1D AAB progression, are ideally needed to establish robust definitions for genetic analyses.

Overall, our results reveal broad differences in genetic risk of T1D in individuals who carry HLA DR3 or DR4 alleles, which supports recent evidence that T1D is a heterogeneous disease with multiple paths to disease. Furthermore, this argues for the need for tailored therapeutic approaches based on genetic background, for example drugs targeting the innate immune response in DR3-T1D.

## Data availability

Summary statistics from genome-wide association studies will be made available in the GWAS catalogue upon publication. Data availability for previous T1D case and cohorts is provided through dbGAP and EGA. Exeter cohort clinical data can be used to identify individuals and are therefore available only through collaboration to experienced teams working on approved studies examining the mechanisms, cause, diagnosis, and treatment of diabetes and other β-cell disorders. Requests for collaboration will be considered by a steering committee after an application to the Genetic Beta Cell Research Bank (https://www.diabetesgenes.org/current-research/genetic-beta-cell-research-bank/). Contact by e-mail should be directed to R.A.O. All requests for access to data will be responded to within 14 days.

## Author contributions

AML and CM performed genetic and genomic analyses and wrote and reviewed the manuscript. KM, ENG, RNB, and GH performed genetic and genomic analyses. WAH, SSR and MNW contributed to study design and reviewed the manuscript. SR contributed resources and reviewed the manuscript. KJG and RAO conceived the study, obtained funding, supervised the study, and wrote and reviewed the manuscript.

## Supplemental Figures

**Supplementary Figure 1.**
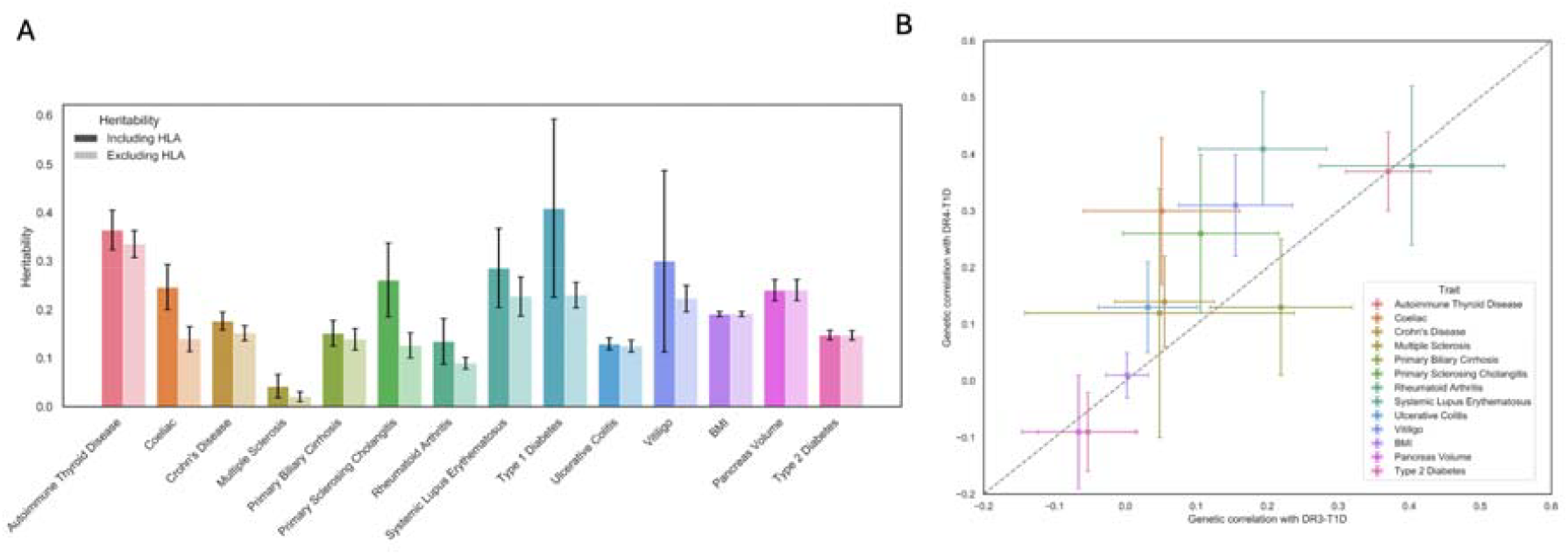
Heritability and Genetic Correlation with DR3-T1D and DR4-T1D. (A) Heritability estimates for autoimmune traits and additional polygenic traits. Darker colors represent estimate with HLA variants, lighter colors exclude HLA variants from estimates. Error bars are standard errors. (B) Genetic correlation of traits with DR3-T1D and DR4-T1D. Dashed line represents equal genetic correlation to DR3-T1D and DR4-T1D. Error bars are 95% confidence intervals.

**Supplementary Figure 2.**
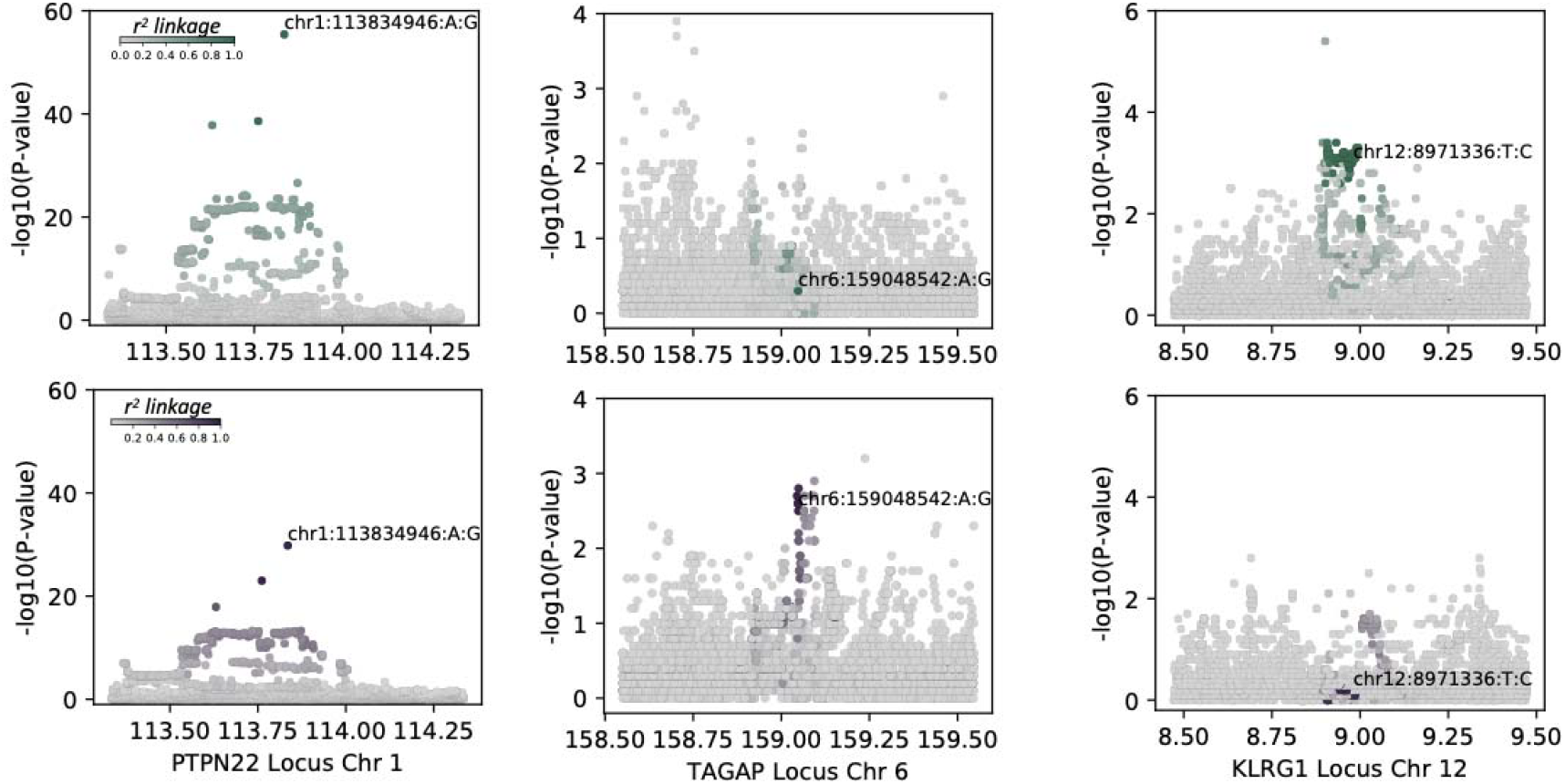
T1D signals with heterogeneity in DR3 and DR4. Locus plots for signals at *PTPN22* (left), *TAGAP* (middle) and *KLRG1* (right) with evidence for heterogeneity between DR3-T1D (green) and DR4-T1D (purple).

**Supplementary Figure 3.**
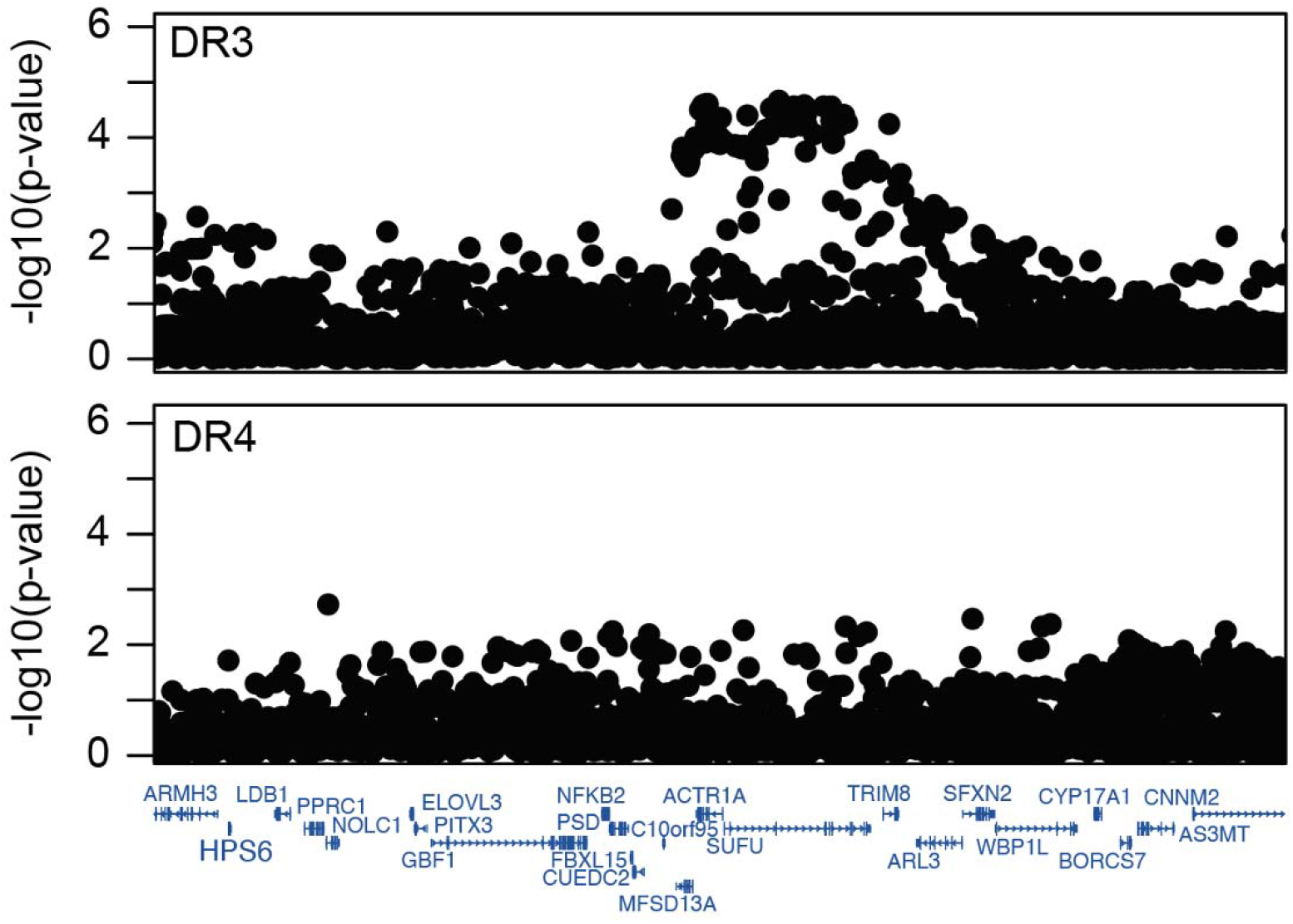
T1D association in DR3 and DR4 at the 10q24 locus. Locus plots showing T1D association of variants in DR3 (top) and DR4 (bottom) individuals at the 10q24 locus.

